# Deep brain stimulation in globus pallidus internus travels to thalamus and subthalamic nuclei along physiological pathways

**DOI:** 10.1101/2023.04.28.23289218

**Authors:** Maral Kasiri, Jessica Vidmark, Estefania Hernandez-Martin, S. Alireza Seyyed Mousavi, Terence D. Sanger

## Abstract

Deep brain stimulation (DBS) is a neuromodulation method for treatment of various neurological disorders. It is often assumed that the primary inhibition or excitation effect of DBS occurs at the site of stimulation. However, recent work has shown that DBS can lead to robust evoked potentials (EP) not only at the stimulation site, representing the local effect, but also in distant brain regions, representing the effects on distant targets. While the significance of these EPs for therapeutic outcomes is not known, it appears that the electrical effects of DBS have a partial modulatory impact on downstream targets. Nonetheless, it remains unclear through what mechanism DBS pulses travel to the distant targets or what portion of the pulses travel along the normal pathways from the stimulation site. The possible scenarios include orthodromic or antidromic pathways, accessory pathways, normally inhibited pathways, and direct electromagnetic activation of distant sites. The ability to record signals from brain regions provides an opportunity to determine the mechanism of DBS signal transmission. We hypothesize that the pathways that transmit DBS pulses include the pathways that transmit intrinsic neural signals. To test this, we performed a transfer function analysis on deep brain recordings during DBS-off condition and compared its impulse response with the transmission of signals from electrical stimulation during DBS-on condition. Our results support our claim that the electrical pulses travel partly along intrinsic neural pathways by showing that the propagation of DBS signals can be partially predicted by observation of intrinsic neural activity and measurement of DBS-EPs.

**New & noteworthy:** This study enhances the understanding of deep brain stimulation (DBS) mechanism by exploring how electrical stimulation travels along neural pathways. We utilized a computational method to explain the main routes through which DBS pulses travel. Our results suggest that DBS signals are likely to be transmitted along the normal pathways. This provides a basis to develop complex and nonlinear models of brain and relate the modulatory effect of stimulation to the brain connectome.

## 1 Introduction

Deep brain stimulation (DBS) is a neuromodulation technique that involves implantation of depth electrodes at potential targets in the brain, through which electrical pulses are administered to modulate neuronal activity [1]. It has been shown that DBS is an effective treatment of various movement and neurological disorders [2], including Parkinson’s disease [3], dystonia [4], essential tremor [5], epilepsy [6], and Alzheimer’s disease [7]. Additionally, recent advancements show that DBS can be used for the treatment of psychological conditions such as obsessive compulsive disorder [8] and major depressive disorder [9]. Despite recent advancements in clinical applications of DBS and its widespread adoption, its underlying mechanism remains poorly understood [4, 10–13]; however, several models have been proposed by researchers on the mechanism of DBS, including the “inhibition hypothesis”, “excitation hypothesis”, and the “disruption hypothesis” [10, 14–16].

Previous research on the mechanism of DBS indicates that DBS effect is similar to those produced by micro-lesions in the brain [17–20], which has led to the replacement of lesion-therapy by DBS [14]. The observed similarity suggested that DBS might inhibit local circuits. Although suppression of neuronal activity in the vicinity of the stimulated region was noted, the “inhibition hypothesis” has been called into question by the detection of DBS-evoked responses (EPs) in distant targets [14].

Other studies have confirmed that GPi-DBS directly induces spiking activity in the GPi neurons, which activates the GABA-ergic (inhibitory) projections onto the thalamic regions. This results in inhibition of those downstream targets, supporting the “excitation hypothesis” of the DBS effect [14, 21–23]. However, this hypothesis was rejected by more recent observations of induced multiphasic responses, consisting of both excitation and inhibition, during GPi-DBS, in the GPi of monkeys with Parkinson’s disease [24–26].

More recently, it has been shown that GPi-DBS during the cortical stimulation inhibits the cortical evoked responses by strong GABAergic inhibition. This suggests that GPi-DBS blocks the information flow through GPi itself, supporting the “disruption hypothesis” [14, 27]. The presented evidences along with other examples [28–30] suggest that DBS essentially blocks the signal transmission from the input to the output of inhibitory or excitatory pathways, resulting in dissociation of input and output [14], rather than having a sole excitatory or inhibitory effect on the downstream regions [14, 27]. This indicates that DBS impact extends beyond its immediate vicinity, with its global influence on distant targets being demonstrated through the recorded EPs. DBS pulses propagate in a specific pattern as evident by the EPs; However, the precise mechanism of propagation and the pulse transmission pathways are not known. Here, we propose a transfer function method to identify what portion of the DBS pulses travel along the neural pathways that carry the intrinsic neural signals.

Identifying the DBS signal pathways is essential for computational modeling and direct measurement of the effects of DBS on brain networks [31]. While anatomic evidence from noninvasive technologies such as diffusion tensor imaging (DTI) is available, this provides only indirect support for anatomical connectivity and is not sufficient for assessing the functional connectivity [32]. Direct measurement is required to learn about the signal transmission along these pathways. Noted that, evidence for the presence of a pathway and the presence of intrinsic signal correlations at either end of that pathway is not sufficient to establish causality or the direction of signal transmission. One way to map DBS pulse transmission in deep brain networks is to stimulate in one region and measure its effect on distant targets [33]. However, electrical stimulation is an un-natural and non-physiological input to the brain. It non-selectively activates and depolarizes a wide group of neurons that would be much more selectively activated in physiology, therefore, essentially, its mechanism of transmission can be very different from that of the intrinsic neural signals. DBS pulses can travel from the source to the target through many pathways, including “normal” (orthodromic) pathways, antidromic pathways, accessory pathways that would not normally be active [11], and direct electromagnetic activation of distant sites (as supported by measuring the volume of tissue activated [VTA] which is partially predictive of the widespread neural effects of stimulation) [34]. Alternative mechanisms also include activation of pathways that are normally inhibited or inactive [14], those with high threshold, or those that are not accessible to stimulation, including polysynaptic pathways [14].

Considering everything mentioned, it has not been studied if the intrinsic signals and the DBS pulses are carried to distant targets through the same mechanism. If they do, what portion of the pathway is directly affected by the DBS pulse, whether it is an orthodromic or antidromic pathway. To distinguish various possibilities, we compared the transmission of DBS pulses to a transfer function representation of intrinsic neural signals (DBS-off local field potentials [LFPs]), at two ends of a known anatomical pathway. The estimated transfer function does not indicate causality, because empirical transfer functions are bi-directional in nature and the causation flow is not clear [35, 36]. Nevertheless, we can set the input and output of the system based on the evidence of physiological connections [14, 37] and extract useful information based on those presumptions. The study methodology was designed to determine whether the normal pathways carry the DBS pulses or the DBS pulses are affecting the distant targets through other mechanisms. We hypothesize that the distant effects of DBS are most likely due to direct transmission of the DBS pulse, perhaps through depolarization of local axons rather than propagation of locally-evoked activity to distant sites.

To test our hypothesis, we made use of intracranial brain recordings (LFP) from Stereoelectroencephalography (sEEG) leads [38], which are used in surgeries for treatment of various neurological disorders, including epilepsy [6, 39], Parkinson’s disease [7], and dystonia [4, 40, 41]. The sEEG leads were implanted into potential DBS targets, as part of the clinical evaluation for implantation of permanent DBS electrodes in deep brain regions of seven children and young adults with dystonia [4, 11, 40]. This was followed by one week of extensive tests and recordings in an inpatient neuromodulation monitoring unit (NMU) with the clinical goal of finding the ideal target region(s) for each patient’s permanent DBS lead(s) [40, 41]. Clinical evaluation focused on capturing EPs in ventral oralis anterior/posterior (VoaVop) and ventral anterior (VA) subnuclei of thalamus, and subthalamic nucleus (STN) during stimulation in globus pallidus internus (GPi), as well as the responses in GPi due to stimulation in VoaVop, VA, and STN [in separate trials]. Clinical data also included LFP recordings during voluntary reaching task, while stimulation was off (DBS-off).

If a significant fraction of the variance of the EPs from the DBS pulses can be explained by a transfer function computed using the intrinsic neural signals (DBS-off recordings), we can claim that DBS pulses may travel via the physiologically used intrinsic neural signal pathways, perhaps through the same mechanism. Additionally, if that is the case, we hypothesize that the fraction of variance in EP explained by the response of the estimated transfer function should be significantly higher in the orthodromic direction than in the antidromic direction, as the intrinsic signals do not travel antidromically. This further supports our hypothesis that the stimulation pulses are primarily transmitted by a mechanism similar to that of intrinsic neural signals.

## 2 Materials and methods

### 2.1 Patient selection and ethics consideration

Seven children and young adults with dystonia (acquired, genetic, or neurometabolic) were selected for DBS surgery if there existed potential stimulation target(s) identifiable with magnetic resonance imaging (MRI) and if alternative medical therapies had been ineffective [42]. The patients were diagnosed with dystonia by a pediatric movement disorder physician (T.D.S.) based on the standard criteria [43]. The patients’ demographics are described in Table 1. Patients or parents of minor patients provided Health Insurance Portability and Accountability Act (HIPAA) authorization for the research use of protected health information and written informed consent for surgical procedures conforming to standard hospital practice, and for research use of electrophysiological data prior to the procedure. The institutional review board of Children’s Health Orange County (CHOC) approved the research use of data and all the surgical procedures and clinical management took place at CHOC, in accordance with standard hospital procedures and policies.

**Table 1:** Patient demographics. All participants are 9-24 year old males.: EP from GPi stimulation exists; ×: EP was not observed due to stimulation in GPi. GA1: Glutaric Aciduria type 1. CP: Cerebral Palsy. *: The GPi-EP exists in all patients due to stimulation in this region.

| Subject | Diagnosis and clinical symptoms | VoaVop* | VA* | STN* |
| --- | --- | --- | --- | --- |
| S1 | CP; Hyperkinetic dystonia | ✓ | ✓ | ✓ |
| S2 | Mitochondrial disease; Hypertonic dystonia, dyskinesias | ✓ | ✓ | ✓ |
| S3 | Mitochondrial disease; Hypertonic and hyperkinetic dystonia | ✓ | × | ✓ |
| S4 | Unknown; Tremor, Ataxia; hyperkinetic dystonia | ✓ | × | ✓ |
| S5 | KMT2B [44]; Chorea, hyperkinetic dystonia | ✓ | ✓ | ✓ |
| S6 | CP; Tremor, hyperkinetic dystonia | ✓ | ✓ | ✓ |
| S7 | GA1; Hypertonic and hyperkinetic dystonia | ✓ | ✓ | ✓ |

### 2.2 Data acquisition

Up to 12 temporary sEEG depth leads (AdTech MM16C; AdTech Medical Instrument Corp., Oak Creek, WI, USA) that are approved for clinical use by the US Food and Drug Administration (FDA) were implanted into potential DBS targets, using standard stereotactic procedure [41, 45]. Based on prior studies of clinical efficacy of DBS in children with dystonia, typical stimulation targets are STN and GPi in basal ganglia, ventral intermediate (VIM), VoaVop, and VA thalamic subnuclei [4, 11, 40]. Each implanted lead contains 6 low-impedance (1–2 *kΩ*) ring macro-contacts with 2 mm height spaced at 5 mm along the lead, as well as 10 high-impedance (70–90 *kΩ*) microwire electrodes (50-µm diameter) referred to as micro-contacts. The micro-contacts are aligned in groups of 2 or 3, evenly spaced around the circumference of the lead shaft, halfway between adjacent pairs of the first macro-contacts. The leads were connected to Adtech Cabrio™ connectors containing a custom unity-gain preamplifier for each micro-contact to reduce noise and motion artifacts. Macro-contacts bypassed the preamplifiers to allow for external electrical stimulation. This setup enables clinicians to stimulate through macro-contacts, and record simultaneously through all contacts. In order to primarily evaluate responses to monosynaptic transmission, we only used micro-recordings from GPi, STN, VoaVop, sampled at 24 kHz by a system with a PZ5M 256-channel digitizer and RZ2 processor, and stored in a RS4 high speed data storage (Tucker-Davis Technologies Inc., Alachua, FL, USA).

The lead and electrode locations were confirmed in a post-hoc analysis, by co-registration of the preoperative MRI with postoperative computed tomography (CT) scan [46]. The lead localization is an important step in the data processing as it provides the exact location of each macro and micro -contacts on the leads within the targeted region. In other words, it provides precise information on the stimulation and recording locations. Figure 1a shows front view of the DBS leads and the segmentation of regions in one patient.

**Fig. 1:**
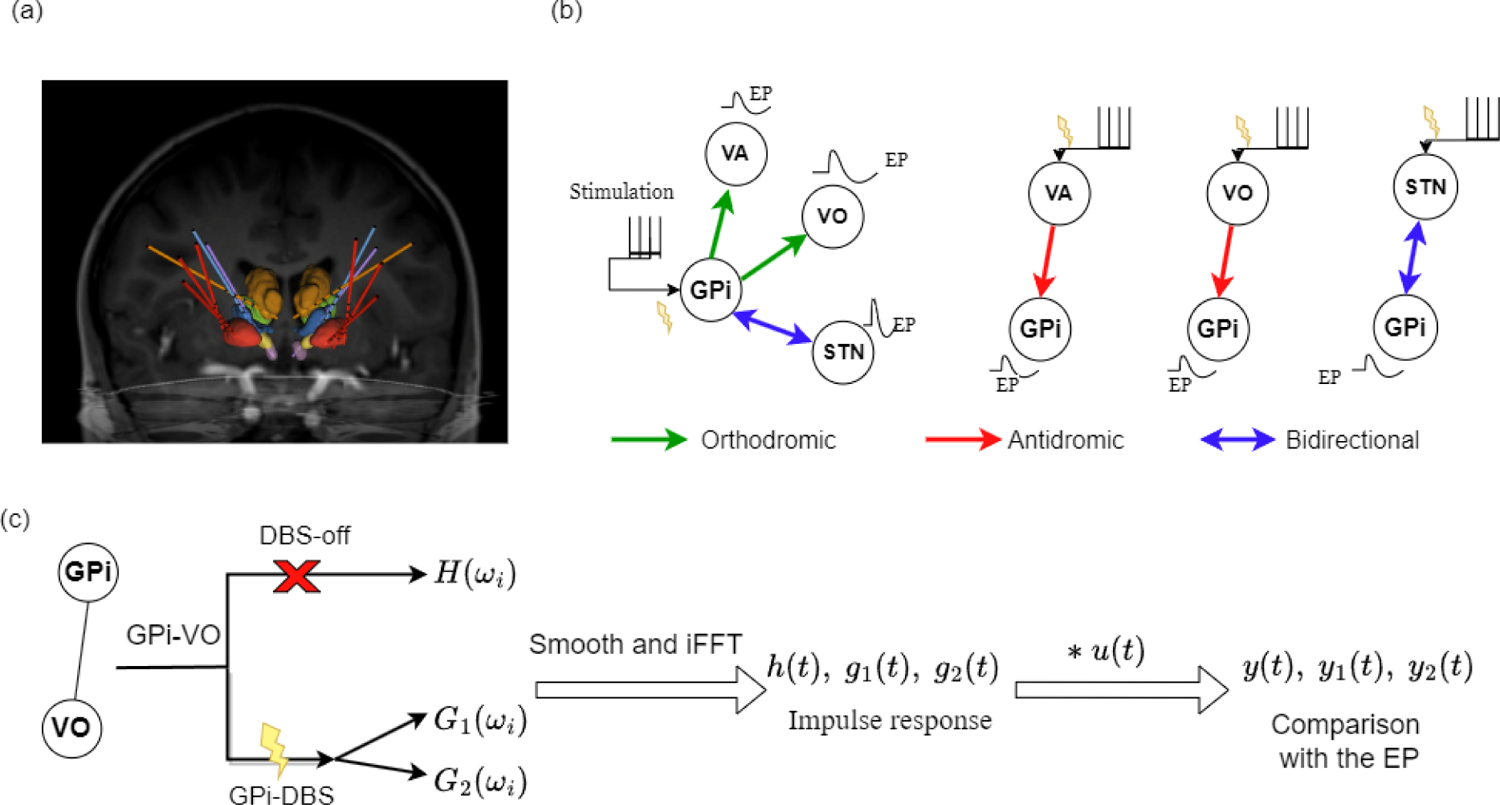
**a)** Frontal view of the DBS leads and the segmented regions in one patient; b) Schematic of all recording and stimulation targets when stimulation is administered in one target, separately. This is a simplified illustration of the pathways and EPs that we used for our analysis and does not imply the precise pattern of EPs due to stimulation; c) Highlights the pipeline of transfer functions computations and comparisons for one pathway connecting the GPi and VoaVop (VO) electrodes. Note that GPi to VoaVop is orthodromic and VoaVop to GPi is antidromic and we computed three transfer functions for this pathway, *H*(*ω_i_*), *G*_1_(*ω_i_*), and *G*_2_(*ω_i_*), in each direction and performed pairwise comparison between their responses, *y*(*t*), *y*_1_(*t*), and *y*_2_(*t*).

The activity in GPi, STN, VoaVop, and VA were simultaneously recorded within 24 to 96 hours after surgery in two modes: 1) While the stimulation was off, and the patient was performing voluntary reaching task with the upper limb contralateral to LFP recordings (intrinsic neural signals; DBS-off condition). 2) while the patient was at rest and unilateral stimulation was on during stimulation in GPi, VA, VoaVop, or STN, in separate trials (DBS-on condition). Intrinsic activity was recorded at rest in order to reduce biasing brain activity toward the movements for which DBS treatment was being tested. Approximately 1200 DBS pulses were administered through two adjacent macro-contacts (anode and cathode) at a time, with 90-*µs* bandwidth and 3-V voltage at 25 Hz, to each nuclei separately, eliciting DBS evoked potentials [EPs], specified in Table 1. Figure 1b depicts a simplified schematic of the trials and stimulation target (inputs: GPi, VA, VoaVop (VO), or STN) and the recording target (output) for each of the EP recordings used in this study. Please note that the stimulation at these targets elicits EPs in other areas of the brain that are not shown here. For example, STN-DBS activates the lenticular fasciculus through activation of hyper direct pathway and provides direct inputs to the thalamic nuclei [14, 47]. However, in this study we are only considering the efferent and afferent pathways to GPi due to its proven efficacy and importance in improvement of dystonic symptoms in our patient cohort [4] and its direct projection on thalamus and subsequently the motor cortex [48]. Moreover, in order to reduce the stimulation artifacts, we stimulated through each contact pair in two trials, with the cathode and anode switched, both of which are located in the same subnuclei; therefore, stimulating similar population of neurons. When the resulting signals are added, the opposite artifact polarities cancel out while the evoked response polarity is augmented. The experimental protocol, movement-LFP synchronization, and the stimulation protocol are detailed in our previously published works, Kasiri et al. [49], Hernandez-Martin et al. [46], and Vidmark, Hernandez-Martin, and Sanger [42].

### 2.3 Data analysis

All data preprocessing and analysis were done in MATLAB R2021a (The MathWorks, Inc., Natick, MA, USA).

#### Data Preprocessing

All the LFPs were notch filtered at 60 Hz including its first five harmonics. They were then high-pass filtered at 1 Hz, to remove the drift. Adjacent micro-contact recording pairs on each lead were subtracted from each other to capture the voltage difference between them, referred to as bipolar montage, resulting in 8 channels per lead. For example, instead of using micro contacts 1 to 3 separately on each contact row, we used their subtraction (1-2, 1-3, and 2-3). This removes the common noise from the data and reveals the underlying neural activity. The LFPs signals that were recorded during the DBS-off condition were then split into 50 two-second segments. On the other hand, LFPs obtained during stimulation underwent another processing step to eliminate stimulation artifacts, which subsequently leads to detection of EPs.

#### DBS artifact removal

After upsampling the DBS-on bipolar recordings to 120 kHz, the stimulus artifacts peaks were located using the ‘findpeaks’ function in MATLAB. The signals were then split into 11-ms segments starting from 1 ms prior to stimulus onset (stimulus artifact). Outlier segments were labeled and removed from the data if the artifact amplitude was not within their *±*3 standard deviations. All the remaining segments were subsequently aligned through cross-correlation of time-0 artifacts. This resulted in approximately 1000 segments per stimulus location, which were finally averaged to increase signal-to-noise ratio [50]. The stimulus-triggered averaging methodology presented by Sinclair et al. [50] was repeated for both sets of polarity reversed stimulation settings, which were finally aligned to produce a polarity-reversed average with smaller stimulus and decay artifacts [42, 51, 52], as shown in Figure 2^4^.

**Fig. 2:**
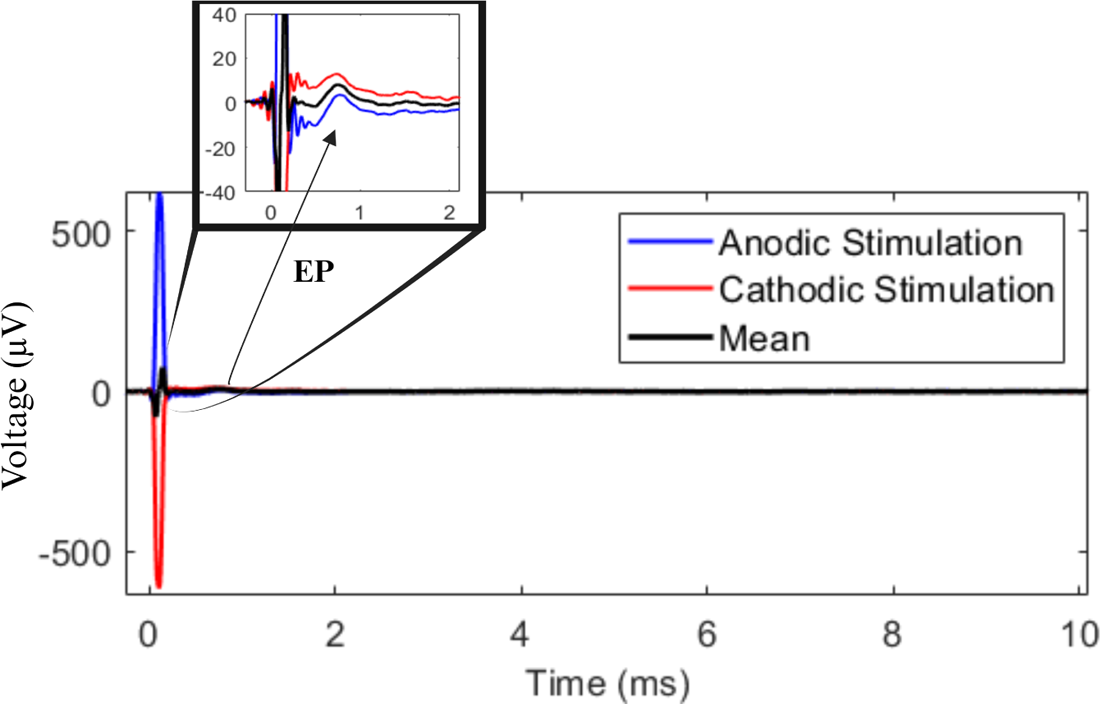
The mean responses of anodic and cathodic stimulation segments are shown in blue and red lines. Their average (black line) cancels out the artifact resulting in smaller decay and stimulus and increasing the signal to noise ratio. The figure is zoomed in along the voltage axis which reveals the actual EP at time *∼* 1*ms*.

#### Transfer Function Computation

In order to determine whether DBS pulses travel via the same pathways that carry intrinsic neural signals, we compared the DBS EPs with the impulse response of a system defined between the same two points in the brain, using intrinsic LFP signals. In other words, we compared the response at a distant region elicited by DBS at the stimulation site with the impulse response of a transfer function between the stimulation site and the distant region in the absence of DBS (DBS-off condition). Transfer functions are specified in the frequency domain, while evoked potentials are specified in the time domain; therefore, we inter-converted the time and frequency domains using Fourier transform (FT) analysis and performed comparisons in time domain [53]. The method pipeline is depicted in Figure 1c.

#### Intrinsic neural signals transfer function computation

We computed the empirical transfer function estimate (ETFE), *H*^°^ (*ω_i_*), from one end of a pathway (input X) to the other end of a pathway (output Y), for each 2-second segment of preprocessed DBS-off intrinsic neural recordings, resulting in 50 distinct DBS-off transfer functions per patient. See Figure 3a for a schematic representation of the system. In this linear time invariant (LTI) system, *H*^°^ (*ω_i_*) is given by

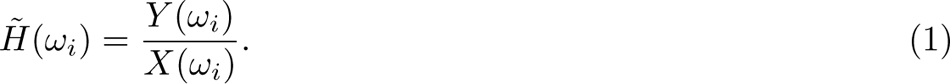

In this equation, *Y* (*ω_i_*) is the FT of the output signal and *X*(*ω_i_*) is FT of the input signal at frequency *ω_i_* and *H*(*ω_i_*) is the transfer function at that frequency [53]. This transfer function is a vector of complex numbers that indicates the gain (amplification) and the phase shift of the input at each frequency, *ω_i_* [53].

**Fig. 3:**
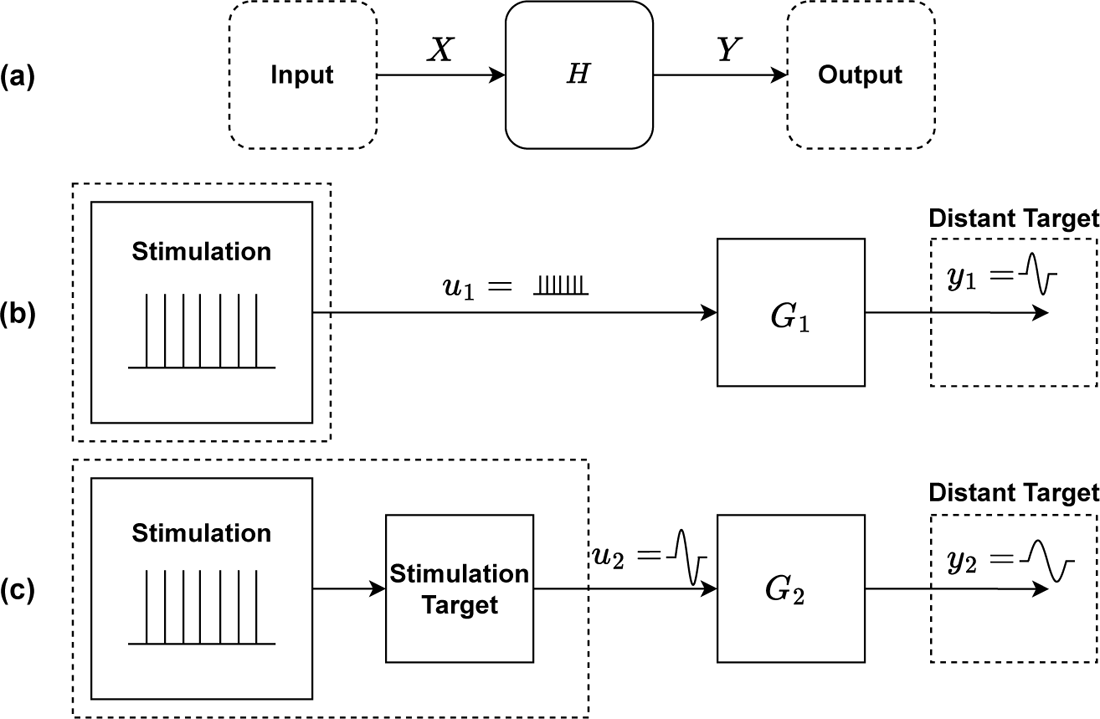
Schematic for the pathways system transfer function: a) Intrinsic neural signal pathway system schematic, where one end of the pathways is the system input and the other end is the system output. b) Direct stimulation of efferent axons by DBS stimulation.; and c) Stimulation of local neurons by DBS, with propagation of the subsequent signal to the target. In case (b), we expect the shape of the DBS signal to be the best predictor of the target response. In case (c), we expect the shape of the local EP at the stimulation site to be the best predictor of the target response.

**Fig. 4:**
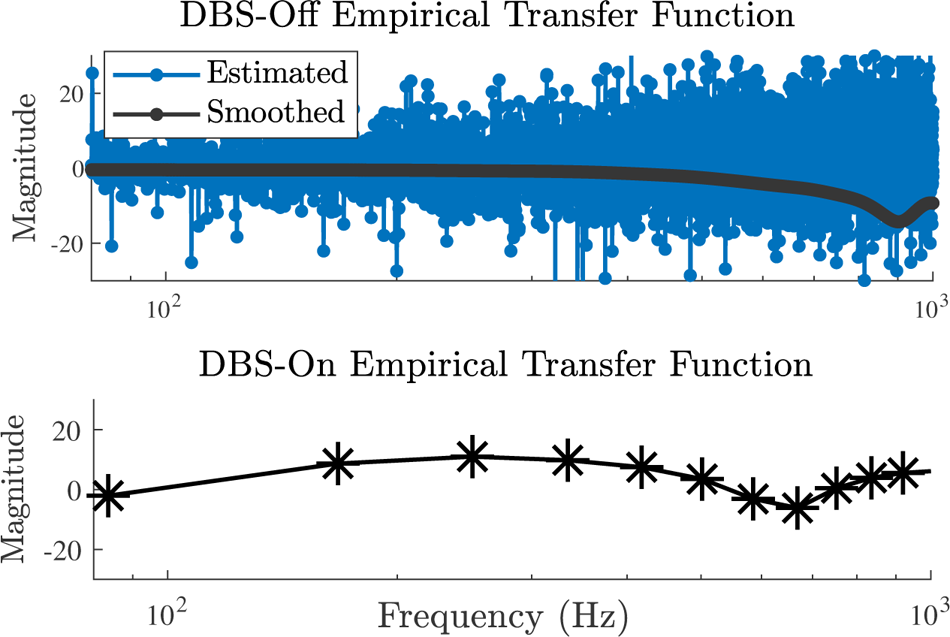
(Top) Bode magnitude plot of a raw and smoothed ETFE from the intrinsic neural signal recordings. (Bottom) Bode magnitude of a stimulation evoked potential transfer function. Note that the smoothing method has no effect on the stimulation evoked potential transfer function, as the response duration is brief (*∼*5 ms) with low number of samples (*∼* 100); confirming that the smoothing method does not introduce distortions to the system.

#### DBS EPs transfer function computation

Using the DBS EPs, we computed two additional transfer functions to investigate whether the target EPs result directly from the DBS signal, or from transmission of local responses of neural tissue near the stimulating electrode. The first case would correspond to DBS depolarization of nearby efferent axons, while the second case would correspond to DBS depolarization of nearby neural cell bodies with subsequent propagation of EP from the stimulation site to the distant target [14, 30, 34, 54]. Therefore, we considered these two cases, in which (a) G_1_ is a transfer function with the stimulation itself as the system input, *u*_1_(*t*), as shown in Figure 3b, and (b) G_2_ is a transfer function with the stimulation-site’s EP as its input, *u*_2_(*t*), as shown in Figure 3c. In both cases, the output is considered to be the distant target’s EP, *y*_1_(*t*) and *y*_2_(*t*). Thus, G_1_ is calculated as the ratio of distant target’s EP FT and the stimulation signal’s FT; and G_2_ is calculated as the ratio of the distant target’s EP FT and the stimulation site’s EP FT.

It is worth noting that although the window sizes for DBS EP and intrinsic LFP signals differ (10 ms versus 2 s), this disparity does not influence the final results. The EP window size represents an average of 1000 segments, resulting in enhanced signal smoothness. We selected a window size of 2 seconds for the intrinsic LFP signals to achieve higher frequency domain resolution and greater clarity when calculating the impulse responses and ETFEs. We then compared the EPs and impulse responses solely for the duration of the response, which is 5 ms. The computed spectral estimates of the intrinsic neural signals (DBS-off condition) transfer functions alter drastically in higher frequencies because the ETFE variance does not diminish with large numbers of samples (Figure 4 top). Therefore, we utilized a smoothing method to smooth out the ETFE [36, 55].

### Transfer function smoothing

Local linear kernel smoothing regression was used to smooth the ETFEs in the frequency domain by solving a weighted least-squares (WLS) problem. The local linear estimator can be obtained by [36, 55–57]:

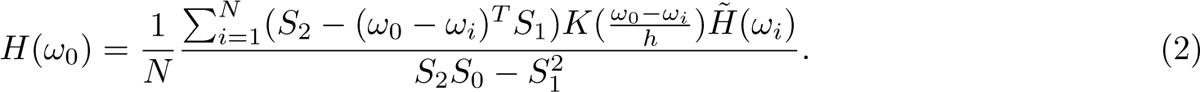

Here, *H*(*ω*_0_) is the smoothed transfer function, *H*^°^ (*ω_i_*) is the non-smoothed transfer function estimate at *ω*_i_, *N* is the number of samples, 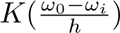 is the Gaussian kernel function, 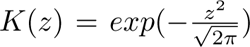, with the bandwidth of *h* = 50 *Hz*, and *S_i_* is given by:

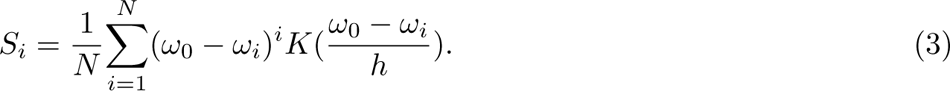

The smoothed ETFE serves as an asymptotically unbiased estimator of the frequency response function [36]. Figure 4 shows an example of Bode magnitude plot of the estimated and the smoothed intrinsic neural signals’ transfer function (top) and an example of *G*_1_ DBS EPs transfer function (bottom), which can be used as a confirmation that the smoothing method does not introduce distortions to the system, since with low number of samples, the smoothed and non-smoothed *G*_1_ transfer functions do not differ significantly from each other.

#### Simulation and Comparison Method

Let *u*(*t*) be a pulse with similar specification to the true stimulation (90-*µ*s pulse width and 3-V amplitude). Let *h*(*t*) be the inverse Fourier transform (iFT) or impulse response of the *H*(*ω*). Thus, the output of the system in time, *y*(*t*), is given by the convolution of *h*(*t*) with *u*(*t*): *y*(*t*) = *h*(*t*) *∗ u*(*t*). We estimated the output of the intrinsic neural signal transfer function (*y*(*t*)) by taking the average of the 50 responses to *u*(*t*) computed for all 50 segments.

Now, let *g*_1_ and *g*_2_ be the iFT of *G*_1_ and *G*_2_, and let *y*_1_ and *y*_2_ be their outputs, respectively. Thus, similarly, *y*_1_(*t*) = *g*_1_(*t*) *∗ u*(*t*) and *y*_2_(*t*) = *g*_2_(*t*) *∗ u*(*t*). Once the responses to *u*(*t*) were estimated, we matched the sampling rates and synchronized all outputs with the respective EPs using cross-correlation. Then, we compared *y*(*t*), *G*_1_, and *G*_2_ responses (*y*_1_ and *y*_2_) with their corresponding EP, in order to first, discover whether *y*_1_(*t*) or *y*_2_(*t*) better replicates the EP, and second, to determine if *y*(*t*) can explain a significant fraction of variance in the EP.

#### Statistical Analysis

First, we compared *y*_1_ and *y*_2_ with the actual EP to determine if the stimulation is transmitted through activation of nearby axons (*G*_1_) or by activation of nearby cell bodies (*G*_2_). Therefore, we computed the fraction of variance explained in EP by *y*_1_ and *y*_2_, and estimated which one is more likely. Then, we computed the fraction of variance explained in EP by the average response of all 50 DBS-off transfer functions computed from the 2-second segments (*y*(*t*)), *R*^2^. This allowed us to verify whether the estimated transfer function from DBS-off intrinsic neural signal data (*H*(*ω*)) estimates the direct EP. We used 50 repeated measures of *R*^2^s for each pathway and each direction per subject to perform the statistical analysis. Among these 50 predictions, we marked the ones that were greater than 3 standard deviation from the mean as outliers (limiting to a maximum of 5 outliers for every 50 segments). Once the outliers were removed, a linear mixed effect model with repeated measure was employed using lme4 [58] package in R-studio (R core team, 2021), with the target nuclei (VoaVop, VA, and STN) as the fixed effect and random intercepts for all subjects. Thereafter, we performed a pairwise multiple comparison using Kenward-Roger’s F-test with the emmeans [59] package to find the differences between each pathway (GPi-VoaVop, GPi-VA, and GPi-STN) by comparison of estimated marginal means, and to discover which pathway response is more likely to be predicted by a linear transfer function representation of that pathway. We adjusted the p-values using the Bonferroni method. The analyses were done with outliers included and outliers removed to confirm that the removal of outliers does not significantly affect the final results.

## 3 Results

### 3.1 Stimulation effect on distant targets: Activation of efferent axons or cell bodies?

For all subjects and all regions, the results showed that *y*_1_ is highly correlated (*R*^2^ = 0.99) with the actual recorded EP. This was expected since *y*_1_ is the result of the convolving the actual EP with an impulse similar to the actual stimulation pulse. On the other hand the *y*_2_ does not have a significant correlation with the actual EP as shown in Figure 5 by the red lines. This provides evidence that the stimulation itself is more likely to cause the EP at the distant target, perhaps by direct excitation of efferent axons near the stimulation site, as opposed to an indirect response due to local cell body excitation. This shows the similarity of *y*_1_ and the actual EP and the inability of *y*_2_ to replicate the EP. Therefore, for the remainder of this paper, EP and *y*_1_ will be used interchangeably, depicted with blue lines in Figure 5.

**Fig. 5:**
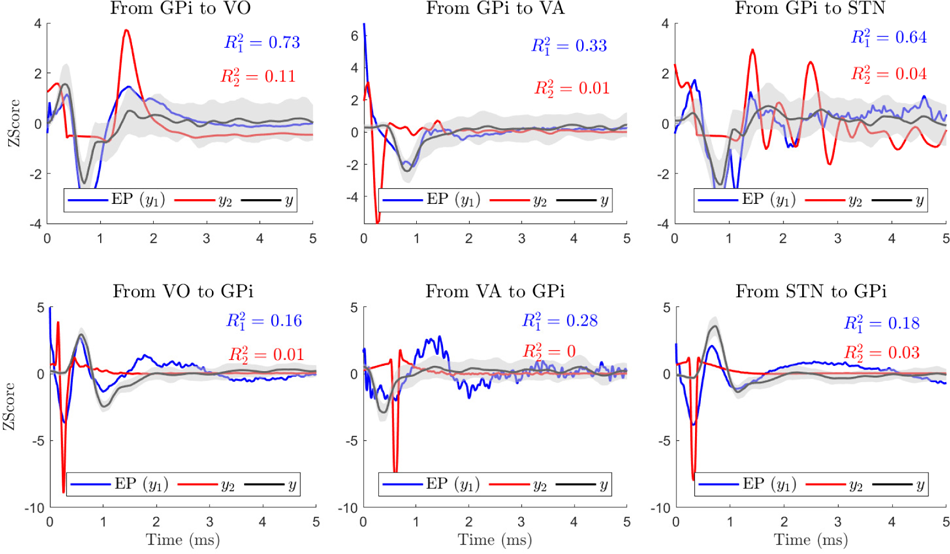
The plots show the true responses in VoaVop, VA, and STN, evoked by stimulation in GPi (*y*_1_(*t*), blue lines). They also illustrate the estimated output of the transfer function *G*_2_ (*y*_2_(*t*), red lines) as well as the response of the intrinsic neural signal transfer function *H* (*y*(*t*), black lines, averaged over *∼*50 segments) and its standard deviation with 95% confidence interval (gray shade). In all cases, the impulse response of the ETFE explains a high variance of direct EP or *y*_1_(*t*), and not the *y*_2_(*t*).

### 3.2 Do DBS Pulses propagate through pathways that transmit intrinsic neural signals?

In order to determine whether the DBS pulses are reaching the distant targets through pathways similar to the intrinsic neural signals or not, we tested the reliability and accuracy of the ETFE impulse responses and compared the predictions for all three pathways in both directions (forward and backward). In order to do so, we only compared our predictions with the EP (*y*_1_(*t*)) for each pathway in each direction by computing the fraction of variance explained in EP by *y*(*t*) (*R*^2^).

The results from comparison of *y*(*t*) with *y*_1_(*t*) (EP) and *y*_2_(*t*) for one subject are shown in Figure 5. As illustrated, the fraction of variance explained in *y*(*t*) by *y*_2_(*t*) (*R*^2^) is not significant in any of the models.

However, the fraction of variance explained in *y*(*t*) (EP) by *y*_1_(*t*) (*R*^2^) is significant in all the models. This result was consistent among all subjects and pathways (GPi-VoaVop, GPi-VA, and GPi-STN), supporting that the EP of the distant target is predicted by an impulse response of the ETFE at the stimulation site, consistent with depolarization of efferent axons.

We used the linear mixed effect model (*R*^2^ = 0.25) fitted to the *R*^2^ (as repeated measures) to compare the quality of prediction in all three pathways, forward and backward. The results of multiple comparison between the pathways, shown in Figure 6, demonstrate that the predicted system outputs have stronger correlation with the DBS EPs in VoaVop and STN compared to the DBS EPs in VA (*p − value ≺.*01). The high fraction of variance of EP explained by the output of the transfer function *H*(*ω*) (*y*(*t*)) reveals that the pathways that transmit external electrical stimulation mostly include pathways that transmit intrinsic neural signals (GPi to VoaVop, GPi to VA, and GPi to STN).

**Fig. 6:**
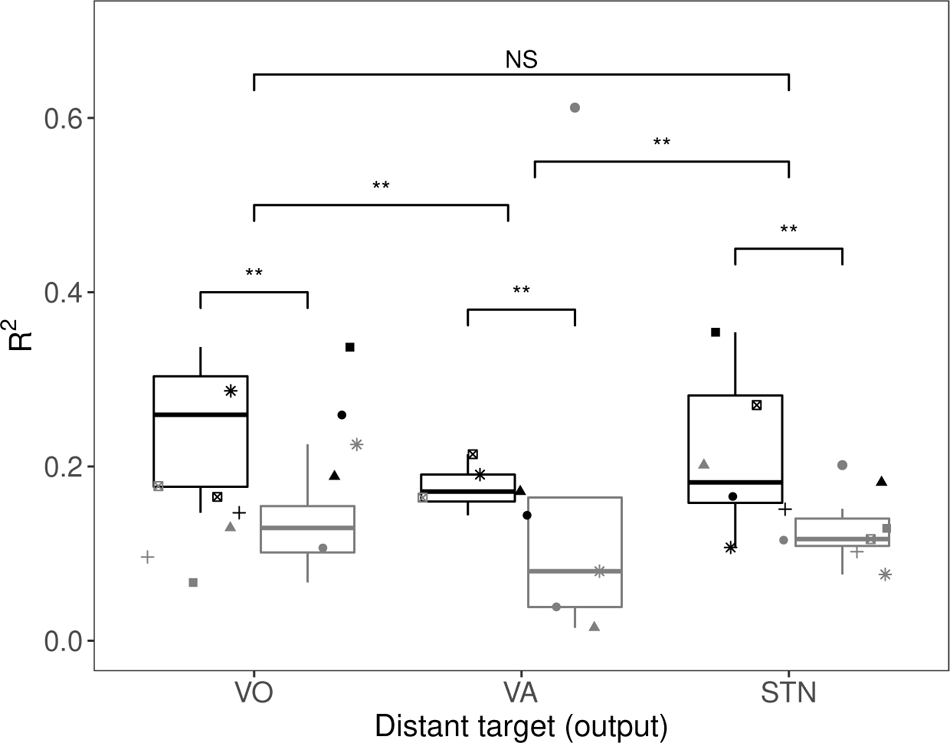
The fraction of variance (*R*^2^) of DBS EP explained by the intrinsic neural signal transfer functions of each pathway (black: pathways from GPi; gray: pathways to GPi) for the seven subjects (each shape represents one subject). The variance explained is greater for STN and VoaVop (*p − value ≺.*01) compared to VA, suggesting that the DBS pulses from GPi to STN and VoaVop are more likely to use the same pathways as intrinsic neural signals, compared to the GPi-VA pathway between the electrode locations. However, the fraction of variance explained in EP by the intrinsic transfer functions of GPi-VoaVop and GPi-STN are not significantly different from each other. Moreover, the results also demonstrate that the ETFEs were able to make better predictions of EP in one direction (from GPi to other targets) compared to the reverse direction (from other nuclei to GPi) (*p − value ≺.*01).

### 3.3 Direction of Signal Transmission

Furthermore, we were able to predict the VA, VO, and STN responses to simulation in GPi significantly better than the GPi response to stimulation in VA, VoaVop, and STN (*p − value ≺.*01), respectively. This observation may be explained by underlying anatomical and physiological differences in the fibers connecting two targets (fiber size, anisotropy, and diffusivity or direction of intrinsic neural signal transmission; ortho-dromic versus antidromic). This also indicates that GPi-thalamus connection is more likely orthodromic, whereas thalamus-GPi is more likely antidromic, or carried by pathways that differ from the normal intrinsic transmission pathways.

## 4 Discussion

In this study we focused on understanding the DBS mechanism of action; as it is significantly important to elucidate the propagation pathways of DBS pulses and to confirm whether these pathways are the same as those utilized by intrinsic neural signals [54, 60]. Previous studies indicate a wide range of potential scenarios for DBS propagation, supported by various models and hypotheses. For example, Zhao et al. [54] demonstrated that STN-DBS in parkinsonian rats activates both motor and non-motor pathways and suggests that this modulation is probably through orthodromic and antidromic pathways. In addition, through an fMRI study on the PD patients, Jet et al. [60] showed that the STN-DBS and VIM-DBS are transmitted to non-stimulated regions through the anatomical pathways, orthodromically and antidromically. However, systematic studies with computational models to explore these scenarios have been rarely conducted. The access to electrophysiological signals and advancements in engineering tools now enable detailed analyses to improve our understanding of the DBS mechanism of action, allowing for a more precise interpretation of how DBS activity propagates, facilitating the development of more optimized and effective approaches for DBS. Here, by using a transfer function analyses, we confirmed that DBS pulses travel at least in part along physiological motor pathways.

### 4.1 DBS affects distant targets through activation of afferent and efferent axons

In the first part of this study, we tested whether DBS pulses directly excite afferent or efferent cell axons near the stimulation site or they excite the efferent cell bodies, through evaluation of EPs due monosynaptic transmission. To achieve this, we compared the EP at a distant target with the empirical impulse response of two transfer functions obtained from DBS-on neurophysiological recordings; one indicating that the DBS affects the distant targets through the direct activation of efferent or afferent axons (*y*_1_) and the other indicating that the DBS effect is through the activation of the nearby cell body, evoking a local response, and its subsequent transmission to distant targets (*y*_2_). Our results indicate that *y*_1_ is nearly identical to the actual EP, which was expected, while *y*_2_ does not explain any variability in the actual EP at all. This result suggests that the DBS pulses are more likely to affect the distant targets through direct activation of the distant area as opposed to the transmission of the stimulation site’s EP to the downstream areas, consequently evoking a response. Here, we are not rejecting other scenarios, but we are providing evidence that the effect of DBS is significantly less probable through the transmission of local EP at the stimulation site to the distant target.

### 4.2 DBS pulse are more likely to travel along the normal anatomical pathways

Next, we compared the DBS EPs with the impulse responses obtained from intrinsic neural signals recorded during DBS-off condition, while patients were performing voluntary reaching movement (*y*(*t*)). If a significant portion of the EP or *y*_1_(*t*) can be explained by the *y*(*t*) we can claim that the DBS pulse and the natural neural activity during movement are transferred to the output through the same mechanism. Our results showed that the fraction of variance predicted in STN, VoaVop, and VA EPs from stimulation in GPi was significantly different from zero, consistent with the hypothesis that DBS stimulation travels at least partly through normal physiological pathways. This provides evidence that the pathways that carry DBS pulses overlap with those that transmit intrinsic neural signals, indicating a similarity in their transmission mechanism.

### 4.3 Orthodromic versus Antidromic Signal Transmission

Previous studies on the mechanism of DBS in Parkinson’s disease demonstrate the complex interaction of DBS and neural fibers in orthodromic and antidromic activation during the stimulation process [47, 54, 61–64]. For example, Kang et al. [62] showed that STN-DBS induced both orthodrimic and antidromic activation through afferent and efferent axonal activation [14] and explored relative contribution of antidromic versus orthodromic effects of STN-DBS in Parkinson’s disease [62].

Here, after we confirmed that the DBS pulses and neural signals pathways overlap, we must validate our method. One way to do it is to perform the same analysis in the direction of antidromic pathways. In this case, we expected to see significantly lower correlation between the DBS-off impulse responses (*y*(*t*)) with the antidromic DBS EPs, since, naturally, intrinsic neural signals do not travel antidromically.

To achieve this goal, we first stimulated in GPi and recorded in thalamus and STN, which makes it reasonable to expect that external evoked responses were primarily carried by orthodromic pathways (specially in VoaVop and VA nuclei of thalamus). Second, we included GPi responses to the thalamic nuclei and STN stimulation in our analyses to compare the ETFE response accuracy in one direction versus the opposite direction. Our results showed that the ETFEs could possibly be a good estimate of direction of pathways in GPi to thalamic subnuclei (VoaVop and VA) projections; orthodromic versus antidromic. The higher correlations of ETFE responses with the thalamic DBS EPs in orthodromic (GPi to VA or VoaVop) versus antidromic (VoaVop or VA to GPi) pathways supports our hypothesis and is a confirmation for use of this method. This suggests that physiological pathways from VA, VoaVop, and STN to GPi are less robust than in the opposite direction, and that a greater fraction of the DBS signal may travel through non-physiological pathways in this direction. However, the higher correlation of GPi-STN impulse responses with the EP versus that of STN-GPi does not reflect the bidirectional connectivity between GPi and STN.

We showed that the GPi-VoaVop and GPi-STN responses have higher correlation with their respective impulse response predictions. The higher correlation of the ETFE responses in GPi-VoaVop, and GPi-STN, (in both directions) with the EPs compared to GPi-VA could be due to the fact that there are fewer projections or fibers connecting GPi to VA, leading to less flow of intrinsic neural signals.

## 5 Limitations

An important weakness of this method is that while the estimated transfer function has an implicit direction, it does not provide evidence of causality, since it is essentially a correlation method [36]. Therefore, the presence of a transfer function from GPi to thalamus does not indicate that activity in GPi is responsible for activity in thalamus. On the other hand, the stimulation results (EPs) do indicate causality, but may only partially correspond to normal physiological transmission pathways. Several other scenarios, including reverse transmission or common drive to both sites, remain possible.

Second potential limitation of this study is that subjects were performing voluntary movement during the recording of intrinsic brain activity, whereas they were at rest during the recording of electrical evoked responses. This element of study design was intentional, in order to evaluate whether signals associated with voluntary (but potentially abnormal) movement flow along the same pathways as DBS responses and whether connectivity in the resting state determines signal flow in the active state. Since we assume that a higher flow of movement-related information in the motor pathways could lead to stronger correlation between two ends of a motor pathway and therefore the obtained transfer function from the intrinsic neural signal is a better representation of that pathway. Moreover, by using this method, we ensure that the correlation between the DBS-off impulse responses and the evoked potential are not related to voluntary movements.

Finally, it is crucial to acknowledge the limitations associated with (linear time invariant) LTI models [65, 66]. LTI models, chosen for their simplicity and interpretability, may not fully capture the dynamic and nonlinear nature of brain function and neural transmission. The brain’s complex and adaptive nature might involve time-varying dynamics that cannot be adequately addressed by LTI models [67, 68]. However, LTI models are powerful tools for explaining the linear behavior of the systems. For example, here, despite the nonlinear nature of the neural activity within the brain, we were able to explain potentially a nonlinear phenomena using a linear method.

## 6 Conclusion and Future Direction

In conclusion, our novel transfer function approach has the potential to describe DBS signal propagation mechanism and possibly, pave the way toward prediction of DBS signal transmission and the causal direction of intrinsic neural signals. Our results imply that electrical stimulation in GPi travels at least in part along pathways that are part of the usual communication between GPi-VoaVop and GPi-STN, and to a lesser extent between GPi-VA. These findings build on what is already established regarding GPi projections to the thalamus nuclei, VoaVop and VA within the motor pathway circuit. This suggests that the natural oscillations dynamics (DBS-off neurophysiological signals) contain useful information about the network responses to DBS pulses, which can be further explored. The results presented here are a first step toward understanding how patterns of therapeutic stimulation interact with the “connectome” to achieve therapeutic benefit. Future work will focus on determining if there are notable differences between anatomical and non-anatomical pathways and studying the impact of DBS on distant brain network underlying activity.

## Data Availability

All data produced in the present study are available upon reasonable request to the authors.

## Acknowledgements

We thank our volunteers and their parents for participating in this study. We also thank Jennifer MacLean for her assistance with neurologic examinations, as well as Jaya Nataraj for helping in data collection. This study is funded by the Cerebral Palsy Alliance Research Foundation (PG02518). Figures 1 and 2 and the graphical abstract are created by biorender.com.

## Footnotes

4 Created with BioRender.com.

